# Comorbid chronic pain and depression: Shared risk factors and differential antidepressant effectiveness

**DOI:** 10.1101/2020.05.23.20110841

**Authors:** William H. Roughan, Adrián I. Campos, Luis M. García-Marín, Gabriel Cuéllar-Partida, Michelle K. Lupton, Ian B. Hickie, Sarah E. Medland, Naomi R. Wray, Enda M. Byrne, Trung Thanh Ngo, Nicholas G. Martin, Miguel E. Rentería

**Affiliations:** Department of Genetics and Computational Biology, QIMR Berghofer Medical Research Institute, Brisbane QLD Australia; Faculty of Medicine, The University of Queensland, Brisbane QLD Australia; UQ Diamantina Institute, The University of Queensland and Translational Research Institute, Brisbane QLD Australia; Brain and Mind Centre, University of Sydney, Camperdown NSW Australia; Institute for Molecular Bioscience, The University of Queensland, Brisbane, QLD Australia; Queensland Brain Institute, The University of Queensland, Brisbane QLD Australia

## Abstract

The bidirectional relationship between depression and chronic pain is well recognized, but their clinical management remains challenging. Here we characterize the shared risk factors and outcomes for their comorbidity in the Australian Genetics of Depression cohort study (N=13,839). Participants completed online questionnaires about chronic pain, psychiatric symptoms, comorbidities, treatment response and general health. Logistic regression models were used to examine the relationship between chronic pain and clinical and demographic factors. Cumulative linked logistic regressions assessed the effect of chronic pain on treatment response for ten different antidepressants. Chronic pain was associated with an increased risk of depression (OR=1.86 [1.37–2.54]), recent suicide attempt (OR=1.88[1.14–3.09]), higher use of tobacco (OR=1.05 [1.02–1.09]) and misuse of painkillers (e.g., opioids; OR=1.31 [1.06–1.62]). Participants with comorbid chronic pain and depression reported fewer functional benefits from antidepressant use and lower benefits from sertraline (OR=0.75[0.68–0.83]), escitalopram (OR=0.75[0.67–0.85]) and venlafaxine (OR=0.78[0.68–0.88]) when compared to participants without chronic pain. Furthermore, participants taking sertraline (OR=0.45[0.30–0.67]), escitalopram (OR=0.45[0.27–0.74]) and citalopram (OR=0.32[0.15–0.67]) specifically for chronic pain (among other indications) reported lower benefits compared to other participants taking these same medications but not for chronic pain. These findings reveal novel insights into the complex relationship between chronic pain and depression. Treatment response analyses indicate differential effectiveness between particular antidepressants and poorer functional outcomes for these comorbid conditions. Further examination is warranted in targeted interventional clinical trials, which also include neuroimaging genetics and pharmacogenomics protocols. This work will advance the delineation of disease risk indicators and novel aetiological pathways for therapeutic intervention in comorbid pain and depression as well as other psychiatric comorbidities.

## INTRODUCTION

Depression is estimated to affect over 264 million people worldwide and is a leading cause of global disability.^1^ Its clinical manifestations and outcomes are highly heterogeneous, with multiple factors underlying susceptibility, progression and treatment response.^2^ One key factor that frequently complicates the diagnosis of depression is comorbid chronic pain, as patients presenting with pain are more likely to be investigated medically rather than as part of a broader biopsychosocial framework.^3^ Depression and chronic pain frequently coexist, with up to 60% of chronic pain patients also presenting with depression.^4,5^ Furthermore, the combination of chronic pain and depression leads to poorer treatment outcomes and overall functioning than either condition alone.^6^ This problem is underlined by depression and chronic pain being among the top three leading causes of global disability over the past three decades.^1^

Chronic pain has been defined by the International Association for the Study of Pain (IASP) as pain persisting or recurring for longer than three months.^7,8^ In contrast to acute pain, which alerts individuals to potential or real tissue damage, chronic pain serves no apparent physiological purpose and persists beyond normal healing time.^7^ In Australia (2015–2016), the disease group with the highest expenditure was musculoskeletal disorders.^9^ In 2016, more than 1.5 million people over the age of 45 had chronic pain^10^ and nearly half of adult patients referred to a pain specialist have comorbid anxiety or depression.^11^ In 2018 the cost to the Australian economy was around $139 billion, mostly due to lost productivity and impaired quality of life^10^ with predictions it will almost triple by 2050.^12^ In the United States, chronic pain is already costing well over $500 billion per year.^13,14^

The relationship between chronic pain and depression is bidirectional, as having either disorder increases the risk of developing the other condition^15–19^ and pain, in particular, is strongly associated with depression onset and relapse.^20–26^ Furthermore, the relationship is dose-dependent with more severe pain being associated with greater severity of depression.^23,27–31^ That is especially true for older age populations which report the highest prevalence (13%)^15^ of comorbid chronic pain and depression out of all age groups.^3,32–34^ However, the evidence for comorbid psychiatric disorders predicting pain intensity and worse outcomes is much weaker.^18,35^ Nevertheless, recent large-scale human genetic studies,^36–52^ animal models^53–56^ and neuroimaging in antidepressant treatment trials^57–59^ have made essential inroads towards delineating the causal mechanisms between chronic pain and depression.

Serotonin noradrenaline reuptake inhibitors (SNRIs; e.g., duloxetine) and selective serotonin reuptake inhibitors (SSRIs; e.g., paroxetine, sertraline) are commonly used antidepressants for the treatment of comorbid chronic pain and depression.^60^ Other antidepressant options include tricyclic antidepressants (TCAs) such as amitriptyline.^61,62^ While these medications have been found to reduce the symptoms of both depression and pain partially, no significant differences in efficacy between them have been established so far,^63^ thus further research is required.^60,64^ For example, the efficacy of TCAs against other antidepressants for the treatment of comorbid chronic pain and depression remains unclear due to a lack of rigorous studies.^65–67^

Despite the high prevalence and cost of comorbid chronic pain and depression,^1,4,5,15–17,19,68–70^ research efforts have yet to deliver clinically useful findings and recommendations specifically for this comorbid indication.^67,71,72^ For example, a recent review highlighted that it was unclear which specific antidepressant should be prescribed as the first-line treatment for comorbid chronic pain and depression,^60^ while others have recommended non-opioid medications as first-line therapy for chronic neuropathic pain.^73,74^ To address this issue, pharmacoepidemiological studies — which examine the use and effect of medications in large population cohorts — have been proposed as a cost-effective method for reviewing pharmaceutical safety and effectiveness, as well as helping to inform clinical guideline development.^75^

In the current study, we examined the pharmacoepidemiology of comorbid chronic pain and depression in the Australian Genetics of Depression Study (AGDS) — one of the world’s largest participant cohorts with a detailed history of depression and its comorbidities.^76^ Here, we sought to: **(i)** quantify the association between depression and chronic pain; **(ii)** assess the dependency between chronic pain severity, depression severity and recent suicidality; **(iii)** identify other psychiatric disorders and patterns of recent substance use associated with comorbid chronic pain and depression; and **(iv)** assess whether comorbid chronic pain and depression is associated with differential antidepressant effectiveness.

## METHODS

### Participants

This study comprised data from two cohorts: AGDS and the Prospective Imaging Study of Aging (PISA). Participants in both groups provided informed consent before participating. These studies, including all questionnaires used, were approved by QIMR Berghofer Medical Research Institute’s Human Research Ethics Committee.

### AGDS cohort

20,689 participants from across Australia were recruited through an open media campaign and targeted mailout. The publicity campaign, from which 86% of participants were recruited, including both conventional and online social media. The campaign appealed for anyone who “***had been treated by a doctor, psychiatrist or psychologist for depression***” to visit this website — https://www.geneticsofdepression.org.au. For the targeted mailout, invitation letters were sent by the Australian Government Department of Human Services (DHS) to individuals who, according to their records, had received at least four prescriptions for any of the ten most commonly used antidepressants in the last 4.5 years. DHS did not, at any time, share any personal information with the research team. Potential participants were directed to the above website, which contained information about the study, a registration and consent form, and a comprehensive online questionnaire. The essential inclusion criteria included having been prescribed and taken antidepressants and providing consent to donate a saliva sample for subsequent genotyping. No participant was excluded based on comorbid conditions. The online survey assessed mental health diagnoses, antidepressant response, suicidality, general health and substance use, among several other variables. A detailed baseline description of the cohort has been published elsewhere.^76^ The full list and details of instruments used for AGDS phenotyping are available at: https://bmjopen.bmj.com/content/bmjopen/10/5/e032580/DC2/embed/inline-supplementary-material-2.pdf

### PISA cohort

The Prospective Imaging Study of Ageing (PISA) is a longitudinal cohort of Australian adults.^77^ The population-based sample recruitment pool comprised adult twins, their spouses, and first-degree relatives of twins and spouses who over previous decades, had volunteered for studies on risk factors or biomarkers for physical or psychiatric conditions and had previously been genome-wide genotyped.^78,79^ The PISA protocol consisted of online questionnaires, including a history of mental health diagnoses and the same pain questionnaire in AGDS. It was completed by N=2,469 PISA participants. For that reason, AGDS and PISA data were used in the present study to assess the effect of depression and demographics (e.g., age, sex) on chronic pain. All other analyses described in this manuscript were performed only in the AGDS cohort.

### Depression and chronic pain ascertainment

AGDS participants were asked to self-report whether they had ever been diagnosed with depression by a health professional, and similarly for 19 other psychiatric conditions. Individuals were classified as depression cases if they had reported both a depression diagnosis and had been prescribed antidepressants in the past five years (N=17,849). Of these, 92% fulfilled the DSM-5 diagnostic criteria for a lifetime depressive episode based on detailed descriptions of this cohort.^76^ Importantly, this figure is within the test-retest reliability estimates of depression ascertainment from DSM-5 based instruments.^80,81^ Participants were administered a pain severity numerical rating scale.^82^ Briefly, patients were asked to indicate whether they experienced chronic pain in their daily life and to rank its intensity on a scale from 0–10. Only those reporting a pain rating >0 progressed to the remainder of the pain module, which included questions about the duration and location of their primary pain. Following the IASP guidelines, chronic pain was defined as pain persisting or recurring for at least three months.^7,8^ Cases were classified as having comorbid chronic pain and depression if they fulfilled the criteria for both conditions (N=6,895), and ***controls*** were classified as those who reported depression but no chronic pain (N=4,475). We performed a complete case analysis. Thus, participants with missing data for chronic pain (i.e., those who did not complete the section; N=6,463) were excluded from analyses that needed data for both chronic pain and depression.

### Recent suicidality and substance use

Suicidality was assessed using the SIDAS instrument.^83^ Briefly, suicidal ideation over the last month was measured on a 10-point scale: 0 indicated having no suicidal ideation in the past month (never), and 10 denoted persistent suicidal ideation. Participants with a score >0 were classified as positive cases for suicidal ideation. Suicide attempt was measured using a similar 10-point scale in regard to how close a participant had come to making an attempt. Only those with a score of 10 (labeled as “***I have made an attempt***”) were considered cases for a suicide attempt. Participants also reported their frequency in using a range of substances over the last three months. Alcohol consumption frequency was measured as the number of days the participant drank three or more standard drinks. For all other substances, the response options were: “***never***” (0), “***once or twice***” (1), “***monthly***” (2), “***weekly***” (3) or “***daily***” (4). These responses were modeled as continuous variables when assessing their correlation with chronic pain.

### Antidepressant use and response

Participants were asked whether they had ever been prescribed any of the ten most commonly used antidepressants in Australia for any indication. These are sertraline, escitalopram, venlafaxine, amitriptyline, mirtazapine, desvenlafaxine, citalopram, fluoxetine, duloxetine and paroxetine. Information regarding the reason(s) for prescription was collected using a checklist of 17 possible responses, including depression, chronic pain, and anxiety (among others). Multiple selections were possible. Participants were asked to report on the best aspects of taking antidepressants using the following item: “***What were the best aspects of taking the antidepressant(s)? Include any antidepressant you have taken***.” Participants were then able to select all that apply out of a list including relief of depressive symptoms, relief of other symptoms, e.g., sleep disturbance, reduction in suicidal ideation, return of normal emotion, improved relationships, returning to normal activities and restored control over mood. Moreover, participants rated the effectiveness of each antidepressant they had taken, using a scale ranging from 0 (e.g., “***sertraline works not at all well for me***”) to 2 (e.g., “***sertraline works very well for me***”). Two analyses were performed: (i) first, antidepressant effectiveness was compared between participants who reported taking an antidepressant prescribed for chronic pain against the rest of the participants (i.e., not prescribed for chronic pain); and (ii) we compared antidepressant effectiveness between participants reporting chronic pain and those reporting no chronic pain (regardless of explicit indication).

### Statistical analyses

In this study, we used complete case analysis and thus removed participants who did not have the required data from specific analyses. The relationship between chronic pain and several other variables of interest was assessed using multivariable logistic regression. This approach enabled us to quantify the associations while adjusting for age, sex and all other relevant factors (e.g., the correlation between alcohol and chronic pain while keeping usage of all other substances ***equal***). Fully adjusted odds ratios were calculated from effect sizes on the logit scale, and p-values were estimated using Wald-tests. For all analyses, the presence of chronic pain was modeled as a binary variable, while chronic pain severity was modeled as a quantitative score from zero to 10. The relationship between chronic pain and antidepressant effectiveness was examined using ***cumulative link logistic regressions*** to accurately model treatment response, which was coded on an ordinal scale. Furthermore, to assess the effect of chronic pain across all antidepressants, a random effect was included to account for repeated responses from participants. This analysis was performed in R using the ordinal package and the ***clm*** and ***clmm*** functions, adjusting for the effects of sex and age when antidepressant treatment started. All other statistical analyses were performed and figures generated in ***python*** using these modules: ***statsmodels, scipy, numpy, pandas, matplotlib*** and ***seaborn***.

## RESULTS

### Sample demographics and association between chronic pain and depression

Demographics and chronic pain prevalence for both AGDS (enriched for depression) and PISA (not enriched for depression) cohorts are shown in **Table 1, Supplementary Figures 1 and 2. Figure 1** shows the prevalence of chronic pain by age, stratified by cohort. A significant cohort effect is evident. Nonetheless, this cohort effect may be attributable (at least in part) to other differences such as age, sex and education rather than depression. Chronic pain is positively associated with age but despite the PISA cohort being older on average (***Supplementary Figure 1***), the AGDS cohort showed a higher prevalence of chronic pain. After adjusting for all the relevant factors, the cohort effect was found to be partly attributable to depression status (OR= 1.86 [1.37–2.54]) because residual cohort effects were non-significant after accounting for the effect of depression (CohortAGDS OR=1.32 [0.97–1.79]). Furthermore, a higher age (OR= 1.02 [1.02–1.03]), lower educational attainment (OR=0.89 [0.86–0.91]), and being female (OR= 1.16 [1.07–1.25]) were associated with chronic pain in the pooled PISA and AGDS sample (***Supplementary Table 1***).

**Table 1.** Chronic pain prevalence in AGDS and PISA cohorts.

| Table 1. Chronic pain prevalence in AGDS and PISA cohorts |  |  |
| --- | --- | --- |
|  | Cases | Controls |
| **AGDS (depression cohort)** |  |  |
| Sample size N (%) | 6,895 (60.6%) | 4,475 (39.4%) |
| Female N (%) | 5,215 (60%) | 3,402 (40%) |
| Age mean (sd)* | 45 (15.1) | 40 (14.3) |
| **PISA** |  |  |
| Sample size N (%) | 1,248 (50%) | 1,221 (50%) |
| Depression* N (%) | 119 (64%) | 68 (36%) |
| Female N (%) | 882 (51%) | 854 (49%) |
| Age mean (sd) | 60 (6.8) | 60 (6.9) |
| Cases: participants reporting chronic pain<br>Controls: participants reporting no chronic pain<br>*p<0.05 two-sample t-test or $\chi^2$ test | | |

**Figure 1.**
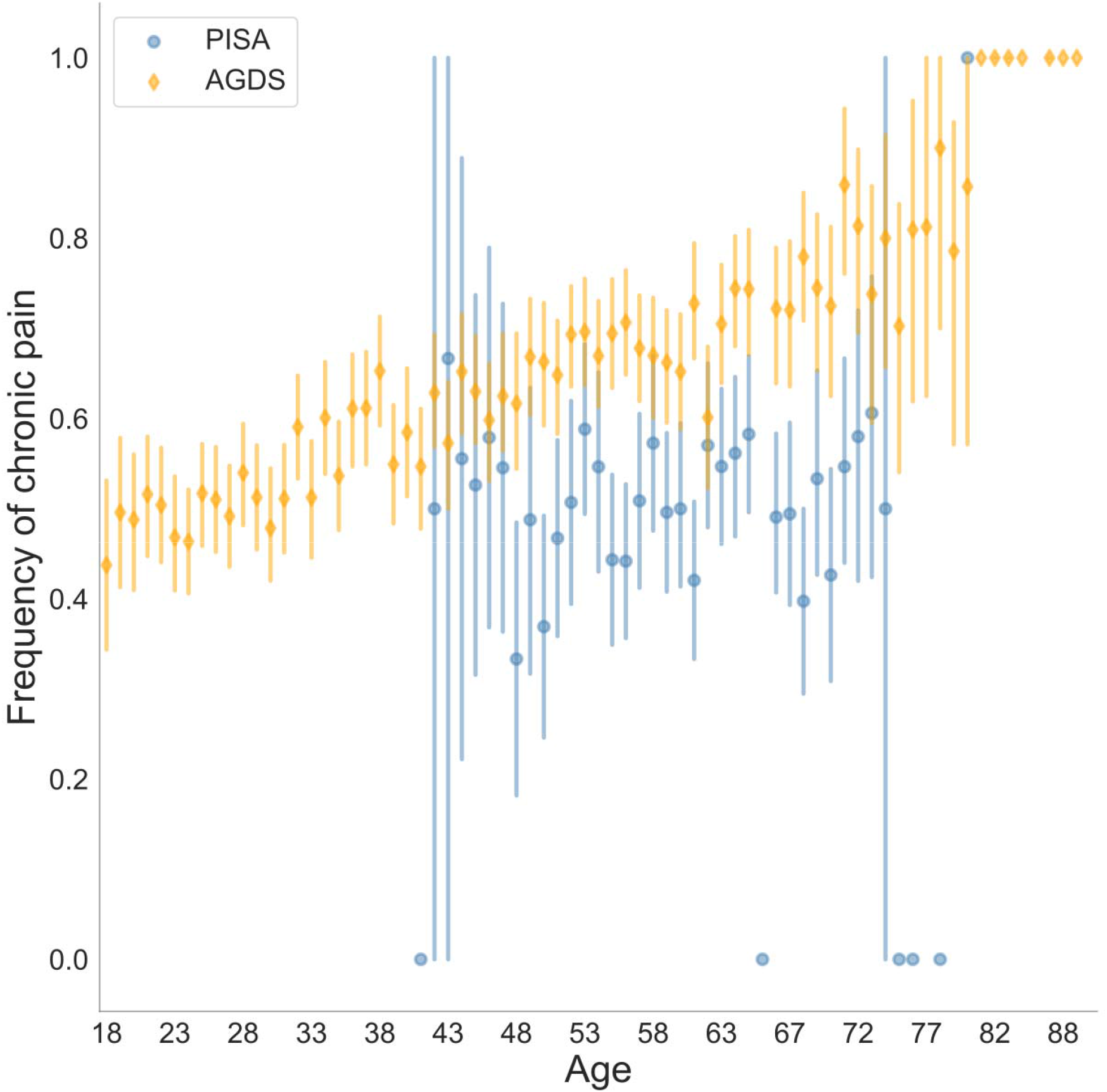
Prevalence of chronic pain stratified by age and cohort (AGDS vs PISA). Self-reported chronic pain was significantly higher in the AGDS cohort (N=6,895/11,370) compared with the PISA cohort (N=1,248/2,469) — OR = 1.31 (0.96–1.77); p=0.086. For other statistical significance results see **Supplementary Table 1**. Both cohorts are population-based samples with AGDS being enriched for depression.

### Chronic pain is associated with severity of depression and recent suicidality

Results presented here are from AGDS where all participants reported depression. Higher pain severity (intensity) was found to be associated with longer durations of pain (***Supplementary Figure 3***). Increased pain severity was also associated with an increased number of depressive episodes (***Supplementary Figure 4***). The prevalence of suicidal ideation was higher in the comorbid chronic pain group (OR=1.49 [1.38–1.61]). Likewise, recent suicide attempt was associated with chronic pain (OR=1.88 [1.14–3.09]). Within the chronic pain group, recent suicidal thoughts and suicide attempt scores were also positively correlated with chronic pain severity scores (***Supplementary Figure 4***).

### Comorbid psychiatric diagnoses and recent substance use

In this subsection, the results presented are from AGDS where all participants reported depression. Out of the nineteen mental health conditions examined, social anxiety disorder was found to have the strongest association with chronic pain (p<0.01), however, this association did not survive multiple-testing correction. Anorexia nervosa was found to be negatively associated with the likelihood of developing chronic pain (p<0.05). Although both of these results were nominally significant, no association survived correction for multiple testing (***Figure 2***; ***Supplementary Table 2***). Notably, chronic pain was significantly associated with decreased use of alcohol, increased use of tobacco, and painkiller misuse (e.g., opioids). Nominal associations were observed for other drugs such as cocaine (negative relationship) and opioids (***Figure 2***; ***Supplementary Table 3***).

**Figure 2.**
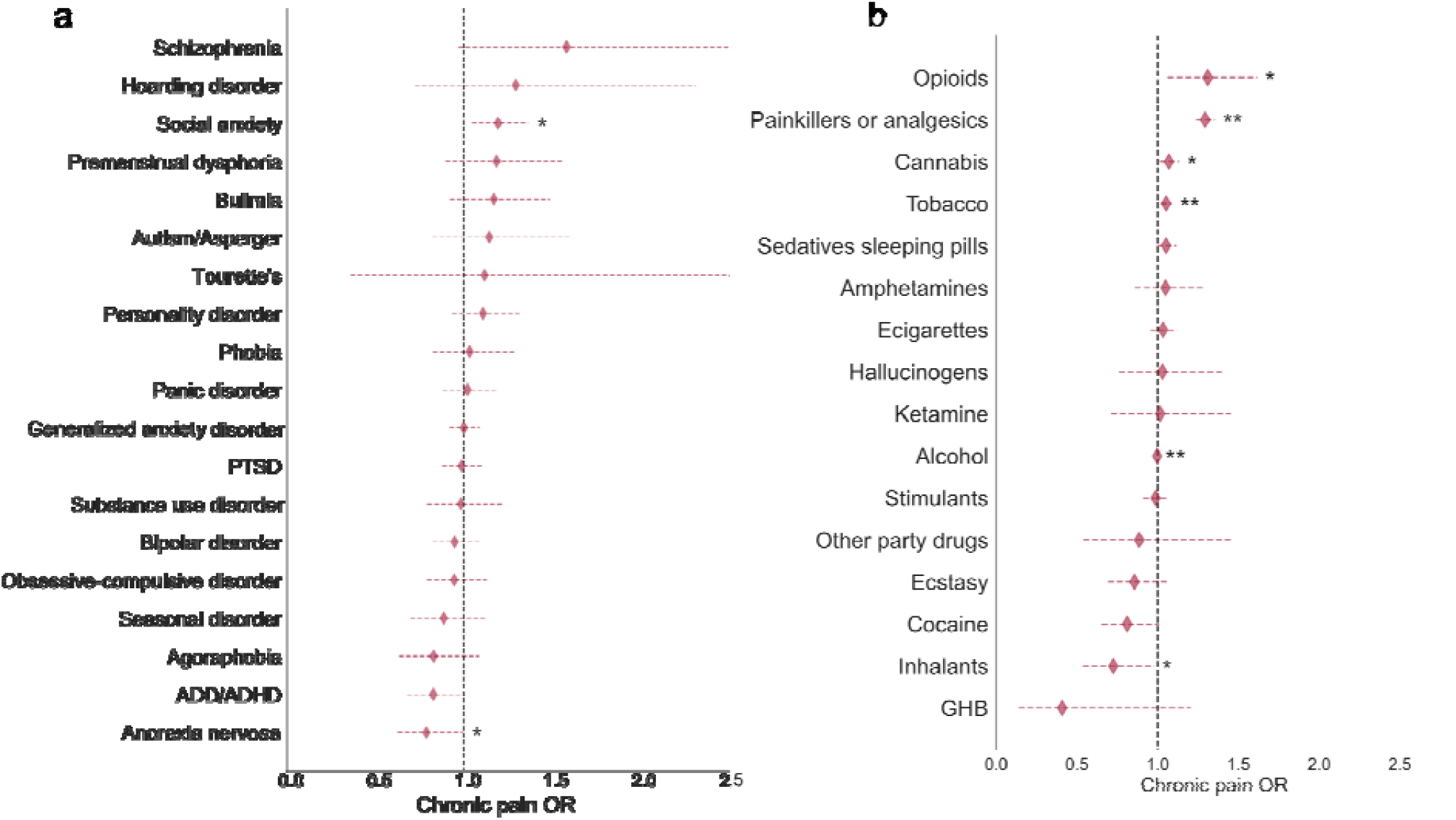
Comorbidities and substance use associations with chronic pain in AGDS cohort. Forest plots depict the chronic pain odds ratios (ORs) for **(a)** comorbid disorders and **(b)** recent substance use during the past three months. Chronic pain was significantly associated with decreased use of alcohol, increased use of tobacco, and painkiller misuse (e.g., opioids). Diamonds represent ORs and horizontal lines depict 95% CI. ORs were estimated from a multivariate logistic regression accounting for all relevant covariates (see Methods). *p<0.05; **p<0.05 after Bonferroni correction for multiple testing (AGDS data only).

### Chronic pain and antidepressant response

In this subsection, the results presented are from AGDS, where all participants reported depression. The three most commonly prescribed antidepressants for the indication of chronic pain — over and above depression — were amitriptyline (N=606), duloxetine (N=288) and sertraline (N=160; **Supplementary Figure 5**). Overall, the overwhelming majority of participants with chronic pain (i.e., 75%–98% across all antidepressants) reported that their antidepressant prescription did not consider chronic pain. Furthermore, participants with chronic pain predominantly reported taking antidepressants for depression (i.e., more than 90% of participants reported their prescription was for depression). The exception was amitriptyline, for which only 60% of participants with chronic pain reported the prescription was for depression (**Supplementary Table 4**). Compared to participants without chronic pain, those with comorbid chronic pain were less likely to report positive functional benefits from taking antidepressants such as ‘***relief of depressive symptoms***’, ‘***return of normal emotions***’ and ‘***getting back to normal daily activities***’ (***Figure 3a***). A trend was noted — whereby participants with chronic pain were more likely to report a ***reduction in suicidal symptoms*** as a positive aspect of antidepressant treatment — but this finding did not survive correction for multiple testing (**Supplementary Table 5**). Furthermore, in the chronic pain group, the average self-reported benefits from taking antidepressants were significantly lower compared to the group without chronic pain (OR=0.75 [0.71–0.80]; p<2×10^−16^). A similar but non-significant finding was observed between the average response of participants prescribed antidepressants for chronic pain versus those without a chronic pain indication (OR=0.94 [0.80–1.1]; **Supplementary Figure 6**). Next, we examined whether these findings held true for each antidepressant under investigation. For most antidepressants, no statistically significant difference in effectiveness was found between participants with chronic pain (or an indication for chronic pain) and participants without chronic pain. Participants with chronic pain who had taken sertraline, escitalopram or venlafaxine, reported significantly lower effectiveness than participants without chronic pain (**Figure 3b**). At this point, it remained unclear whether the antidepressant was taken before or after the commencement of chronic pain. To address this question, we performed a secondary analysis defining cases as participants who reported taking an antidepressant where the prescription explicitly considered chronic pain. This analysis revealed lower effectiveness for sertraline, escitalopram and citalopram(**Figure 3c**). The only antidepressants with a positive effect (i.e., greater effectiveness) were duloxetine, venlafaxine and amitriptyline, but only when prescribed for comorbid chronic pain and depression. However, none of these positive associations reached statistical significance (**Figure 3b**; **Supplementary Table 6**; **Supplementary Table 7**).

**Figure 3.**
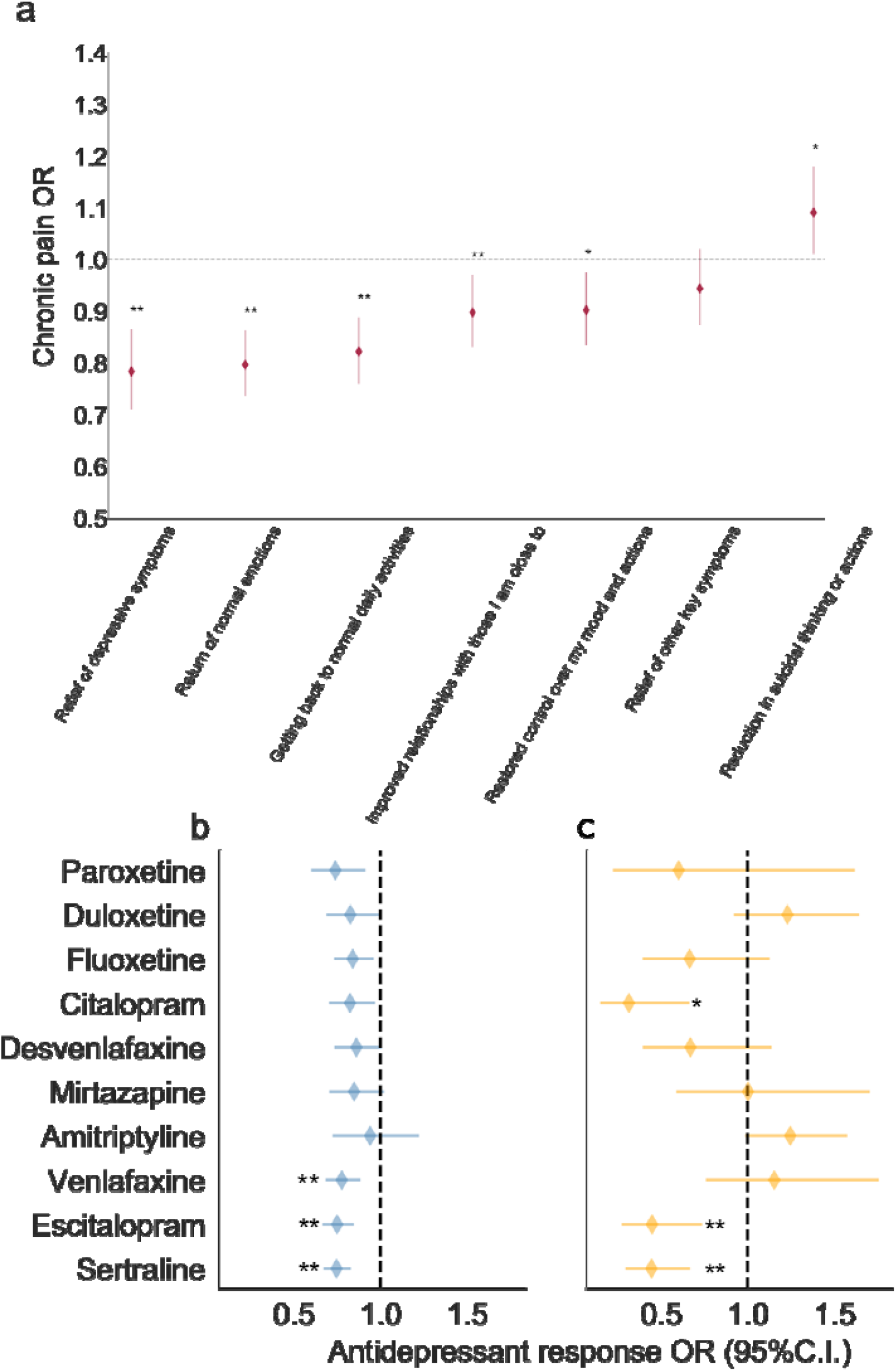
Effect of chronic pain on antidepressant benefits in AGDS cohort. Forest plots depicting the results from: **(a)** association of chronic pain with self-reported benefits from general antidepressant treatment; and associations between antidepressant treatment response and **(b)** self-reported chronic pain or **(c)** self-reported prescription for chronic pain, while adjusting for the effects of sex and age at commencement of taking the antidepressant (*p<0.05; **p<0.005 statistical significance after Bonferroni correction for multiple testing ; AGDS data only). Further details are in Supplementary Figure 4 and Supplementary Tables 4 and 5.

## DISCUSSION

We have reported the largest study to date on comorbid chronic pain and depression assessing the risk factors and treatment outcomes through comprehensive phenotyping of a depression-enriched sample. A key finding is that participants with comorbid chronic pain and depression reported significantly lower benefits from taking particular SSRI and SNRI antidepressants (i.e., sertraline, escitalopram, venlafaxine) compared to participants with depression but no chronic pain. Participants with comorbid chronic pain and depression also reported fewer functional benefits from taking antidepressants compared to those without chronic pain. For example, participants with comorbid chronic pain and depression treated with antidepressants were 22% less likely to report relief of depressive symptoms and 18% less likely to get back to normal daily activities compared to depression patients without comorbid chronic pain. The fewer functional benefits reported from antidepressants in those with comorbid chronic pain is consistent with prior research showing that pain is a strong predictor of non-remission with antidepressant medication treatment.^84,85^

We also found that participants prescribed particular SSRI antidepressants (i.e., sertraline, escitalopram, citalopram) ***for chronic pain*** reported significantly lower benefits (e.g., 55% lower odds of response from sertraline) compared to those taking the same medications but for a different indication. These results suggest that while SSRI and SNRI antidepressant classes may be equally effective in the treatment of comorbid chronic pain and depression,^63,86^ specific antidepressants have differential effectiveness depending on certain common disease modifiers such as chronic pain. The lower effectiveness of sertraline is particularly important, as it is commonly used for the treatment of comorbid chronic pain and depression.^60^ Here we have shown evidence for differential effectiveness between several specific antidepressants in comorbid chronic pain and depression. We consider these findings to be robust because our methodological approach took into account the inherent clinical heterogeneity, high comorbidity and wide individual variation commonly observed in psychiatric disorders.

The current study’s main findings of differential antidepressant effectiveness and fewer functional benefits from antidepressant use in comorbid chronic pain and depression are further underlined by demonstrating several results consistent with previous research. These include: **(i)** a strong association between depression and chronic pain;^87,88^ **(ii)** increasing severity of chronic pain was associated with a higher number of depressive episodes experienced by participants;^23^ and **(iii)** older age, lower educational attainment and female sex were associated with higher chronic pain prevalence.^3,32–34,89–91^

In the current study, amitriptyline was found to be the most commonly prescribed antidepressant to individuals with comorbid chronic pain. Indeed, it was prescribed over two times more than the next most commonly prescribed medication — duloxetine. Moreover, amitriptyline was the medication with, by far, the lowest indication for depression followed by duloxetine. Amitriptyline has traditionally been the first-line treatment for chronic neuropathic pain^61^, however, its side-effect profile and mortality risk in overdose often limit its use.^67^ Our results are consistent with these medications being effective at treating chronic pain. In the current study, when the antidepressant prescription was for chronic pain — amitriptyline, duloxetine and venlafaxine showed a positive association with treatment effectiveness. This effect did not survive multiple-testing correction, which may be explained by the reduced power from further stratifying the sample. Our findings highlight the inadequate treatment recommendations for comorbid chronic pain and depression, as most participants with chronic pain did not report their antidepressant prescriptions were for chronic pain. The current study thus reaffirms the critical unmet need in this patient population.

While there have been conflicting reports regarding the link between chronic pain severity and suicidal behaviours,^92–97^ we provide evidence that supports an association between comorbid chronic pain and depression with both an increased risk for suicidal ideation and suicide attempt. Given suicide is a leading cause of death — particularly for young people^98^ — and that depression and chronic pain are both treatable conditions, assessing their comorbidity in both at-risk youth and older adult populations may help to reduce suicide rates.^99–102^

Consistent with previous observations,^88^ we also found comorbid chronic pain and depression was associated with recent increased use of tobacco and painkillers (e.g., opioids). However, we did not observe a significant association between comorbid chronic pain and depression with a self-reported substance use disorder. Previous reports suggest chronic pain, depression and substance use disorder are often comorbid.^103^ It is possible that screening and diagnosis of substance use disorders in Australia may be lacking in those with comorbid chronic pain and depression. As such, clinicians need to consider substance use disorder in patients presenting with this comorbidity, as all three conditions increase the risk for other chronic diseases such as cardiovascular disease and cancer, while also increasing the risk of premature death.^71^

### Strengths, Limitations & Further Research

The current study is the largest to date examining the relationship between depression and chronic pain with a novel pharmacoepidemiological approach that has yielded new insights into the medical treatment of these highly morbid conditions. However, there are also a number of limitations to be acknowledged. Data were primarily drawn from AGDS, which is a multi-aim study investigating the risk factors for depression and treatment response to antidepressants. As the AGDS employed phenotyping across an extensive range of complex traits^104^, on balance it was not feasible to also collect detailed information from participants on dosages and (polypharmacy) combinations taken of the prescribed antidepressants^105^; the duration and magnitude of benefits (e.g., for cost-effectiveness analyses)^66,106,107^; drug tolerability and adverse events^66,108–113^; adjunct psychological therapies and multidisciplinary treatment/rehabilitation programs;^114^ other prescribed pain pharmacotherapies and questionnaires.^115–119^ Further pharmacoepidemiological studies focusing on chronic pain and a large range of psychiatric comorbidities^51,52,120^ can directly address and collect data on these specific issues. As the current data were based on self-reported responses, they may also be subject to a degree of participant recall bias (e.g., time-specific details). For example, the substance use and suicidality phenotypes may be subject to non-disclosure effects due to the potential stigma associated with these conditions and the antidepressant response data may include non-specific (placebo/nocebo) effects.^59,121^ However, these effects would be present across all antidepressants and thus alone are highly unlikely to explain the observed differences between the chronic pain (cases) and control participants. Randomized interventional studies comparing treatments for participants with comorbid chronic pain and depression are required to validate our results and elucidate the causal direction of associations reported here. Genetic-based methods can also aid in further examination of our findings by performing discovery genome-wide association studies (GWAS) of comorbid chronic pain and depression to determine individuals’ polygenic risk irrespective of whether they have developed chronic pain or not. Furthermore, GWAS will enable: **(i)** the elucidation of causality between chronic pain and treatment response by using methods such as Mendelian randomization; and **(ii)** replication in antidepressant treatment-resistant depression cohorts with primary care and genotype data^122^.

### Conclusions

In summary, we found patients with comorbid depression and chronic pain were less likely to derive functional benefits from antidepressants (especially sertraline, escitalopram and venlafaxine) than patients with depression but no chronic pain. Compared to patients with depression but no chronic pain, those with comorbid chronic pain and depression were also more likely to have had a recent suicide attempt, use tobacco and misuse painkillers. To further assess differences in effectiveness between specific antidepressants, targeted interventional trials can directly address the other phenotypes not captured in the current study. Nevertheless, our large-scale data-driven approach — like recent human genetic^36–52^ and neuroimaging^57–59^ antidepressant treatment studies — have revealed novel insights into the relationship between chronic pain and depression. Along with animal model and human pharmacogenetic studies^53– 56,118,119,123–127^, there is also independent converging evidence for the critical role of subcortical brain regions in mediating pain and mood.^128^ The application of rigorous statistical genetics methodologies to large-scale neuroimaging data, for example, has already produced several major discoveries, such as advancing our understanding of causal pathways, (subcortical) brain networks and medication response markers in mood disorders.^129–135^ Our study suggests pharmacoepidemiological approaches in psychiatry and pain medicine research will be increasingly valuable as a cost-effective, first-line strategy to enhance the design, feasibility and clinical utility of randomized controlled trials^136^, particularly as they routinely exclude patients with specific comorbidities and thus are not representative of the inherent individual variation across the population.^106,109,137^ Finally, the current study also has important implications for Australia’s mental health system and chronic disease policy reforms, such as addressing problems concerning medication overprescription and effectiveness, their side-effects and suicide prevention.^138–140^

## Data Availability

Code used as part of this study is available from the authors upon reasonable request. Participant data is protected and confidential.

## AUTHOR CONTRIBUTIONS

WHR, AIC, TTN and MER designed this study and wrote the first version of the manuscript. AIC performed the analyses with input from MER, TTN and WHR. NGM, SEM, NRW and IBH designed and directed the AGDS data collection efforts. NGM and MKL led the PISA study data collection efforts. TTN designed the pain module in both the AGDS & PISA online surveys and conceived the genetic & epidemiological investigation of comorbid pain & depression in these cohorts. LMG-M and GC-P contributed to data analyses. All authors contributed to the interpretation of the results and provided feedback on the preliminary versions of the manuscript.

## ACKNOWLEDGEMENTS

Data collection for AGDS was possible thanks to funding from the Australian National Health & Medical Research Council (NHMRC) to NGM (GNT1086683). PISA was possible thanks to an NHMRC Dementia Research Team Grant administered by QIMR Berghofer Medical Research Institute (GNT1095227). We thank our colleagues Richard Parker, Simone Cross and Kerrie McAloney for their valuable work coordinating all the administrative and operational aspects of the AGDS and PISA projects. AIC and LMGM are supported by UQ Research Training Scholarships from The University of Queensland (UQ). MER thanks the support of NHMRC and the Australian Research Council (ARC), through an NHMRC-ARC Dementia Research Development Fellowship (GNT1102821). The views expressed are those of the authors and not necessarily those of the affiliated or funding institutions.

## SUPPLEMENTARY FIGURES

**Supplementary Figure 1.**
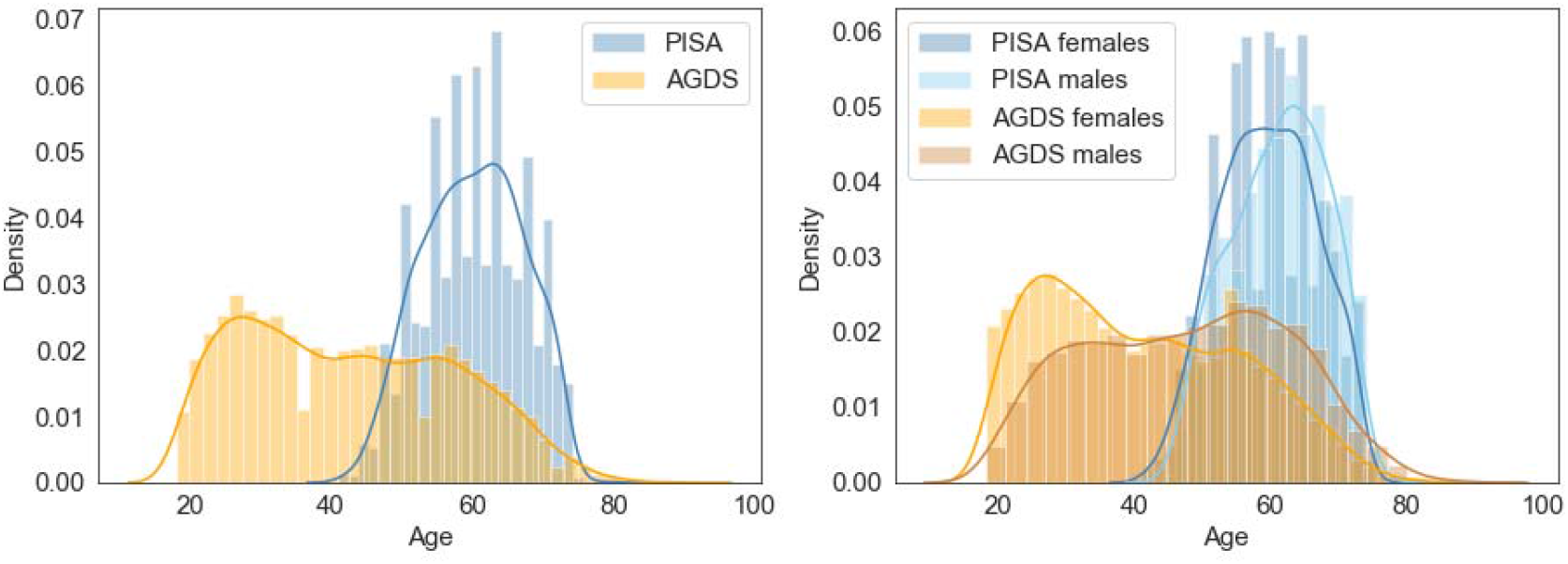
Age and sex distributions in AGDS and PISA cohorts. Histograms with kernel density plots showing (**left**) the distribution of ages for AGDS and PISA cohorts and (**right**) after stratification by sex. Note that the PISA cohort is on average older at the time the participants responded to the questionnaire, but have lower rates of chronic pain on average than the AGDS (depression-enriched) cohort.

**Supplementary Figure 2.**
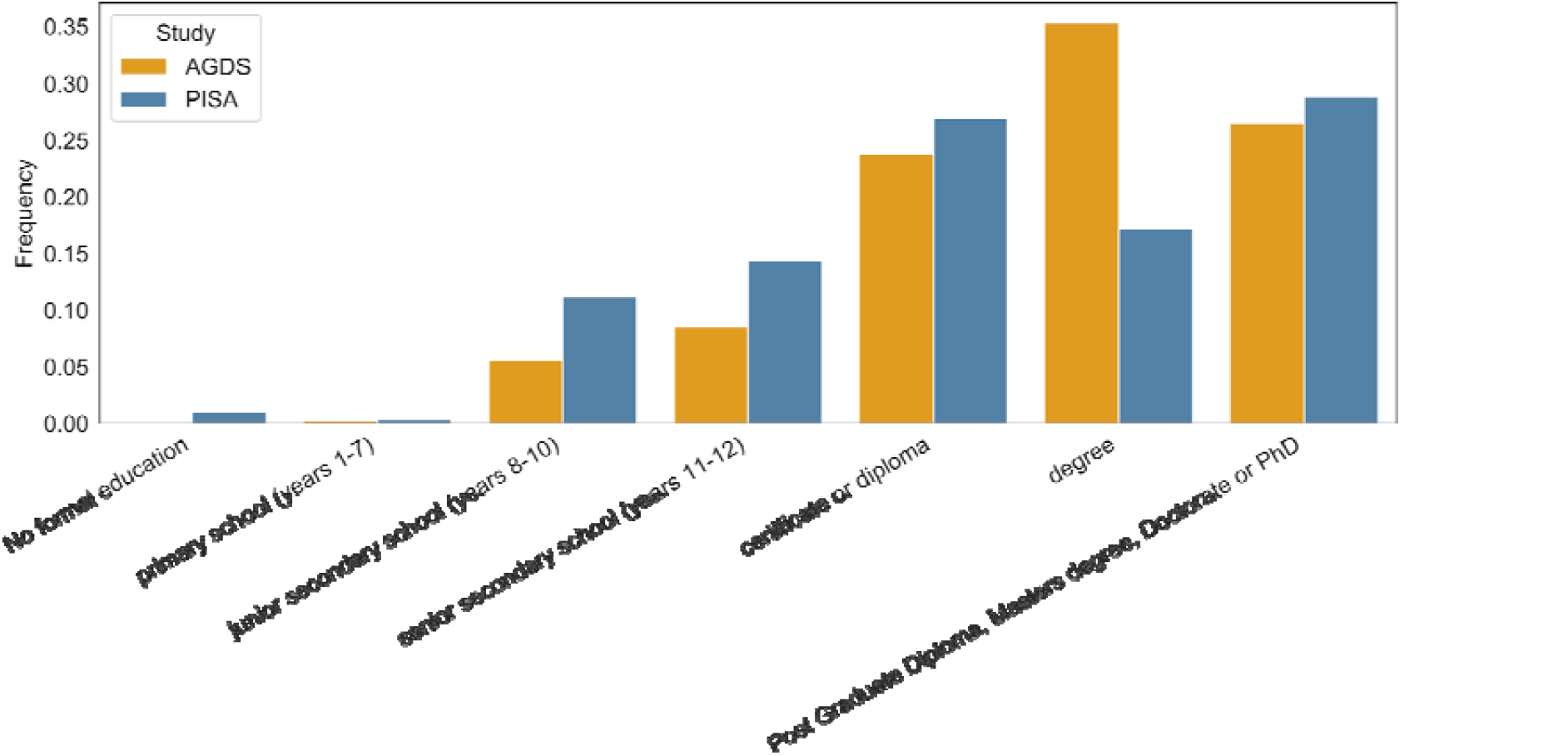
Significant difference in educational attainment between AGDS and PISA cohorts. Barplots showing the frequency of educational attainment for the AGDS and PISA cohorts. The difference is statistically significant (χ2 test p<E-100).

**Supplementary Figure 3.**
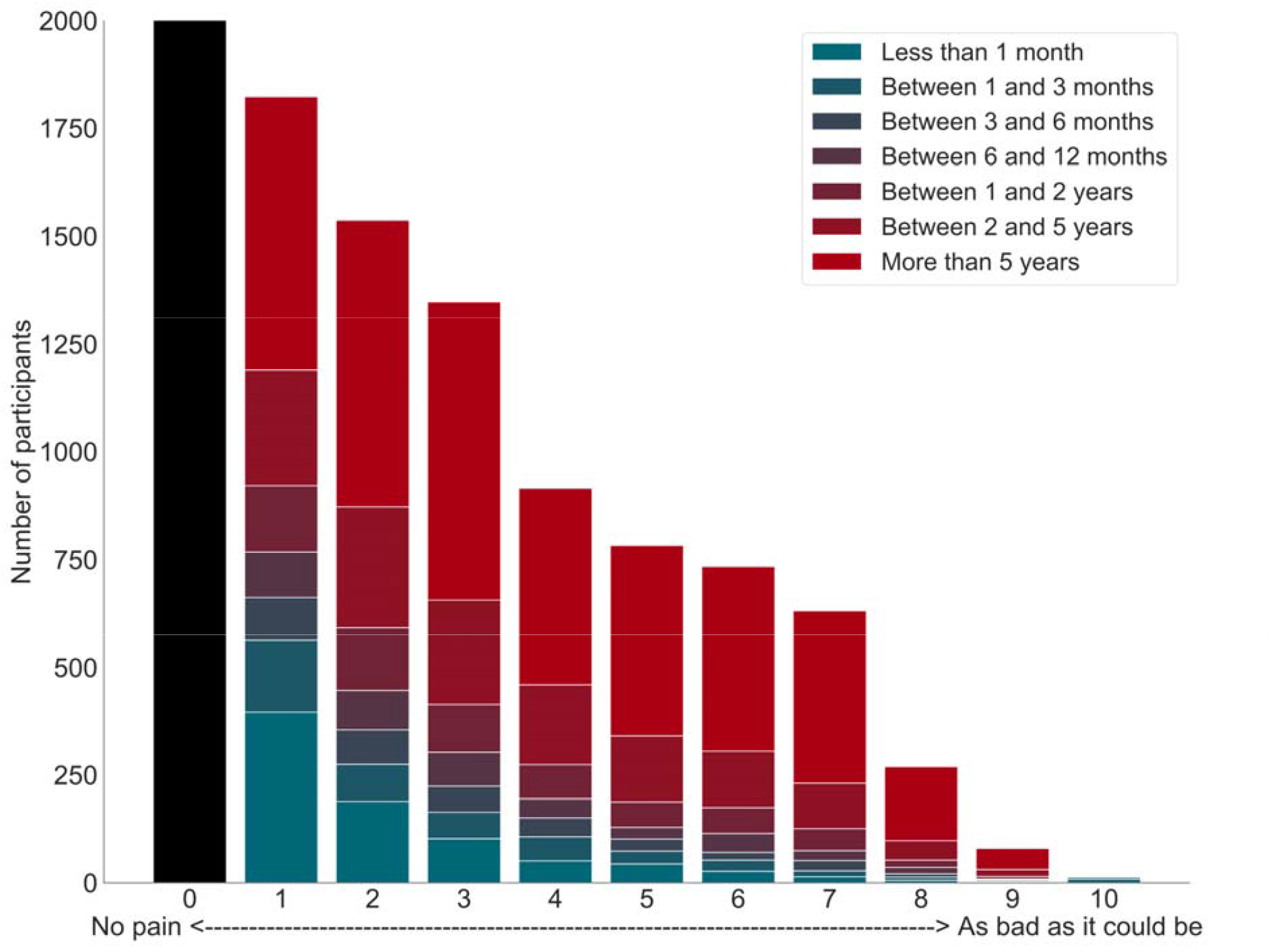
Positive association between pain intensity and pain duration in AGDS cohort. Bar plots illustrate average pain intensity (x-axis) stratified by pain duration, with a strong positive relationship found between these measures (p=5.5e-106 linear regression; AGDS data only).

**Supplementary Figure 4.**
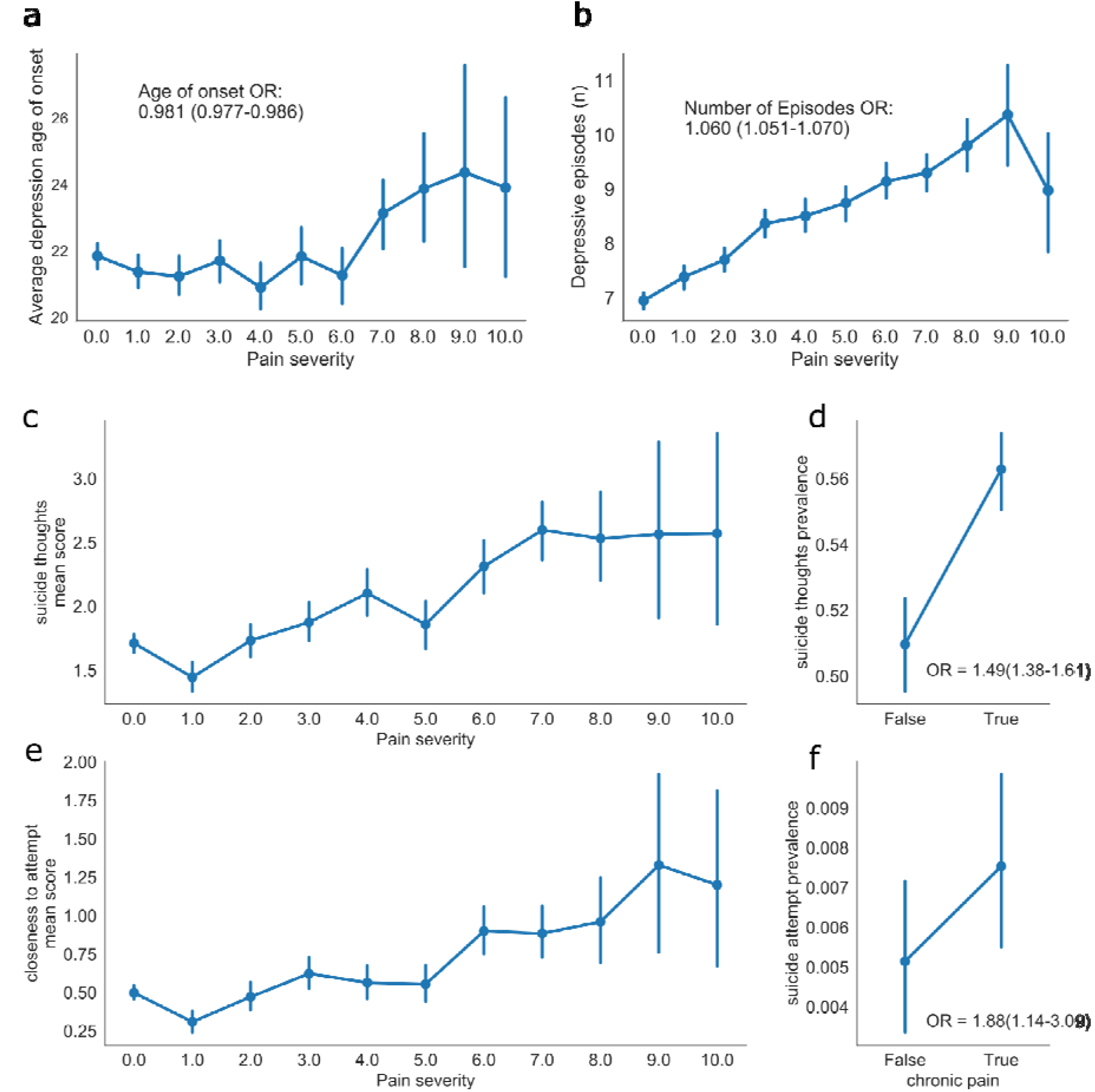
Positive association between chronic pain severity (intensity), depression (number of episodes) and suicidality in AGDS cohort. Point plots comparing chronic pain severity with: **(a)** average age of onset; **(b)** average number of depressive episodes; **(c)** average suicide thoughts score; or **(e)** suicide attempt score. Also shown are differences in the prevalence of **(d)** recent suicide thoughts and **(f)** recent suicide attempts. Error bars denote 95% confidence intervals (AGDS data only).

**Supplementary Figure 5.**
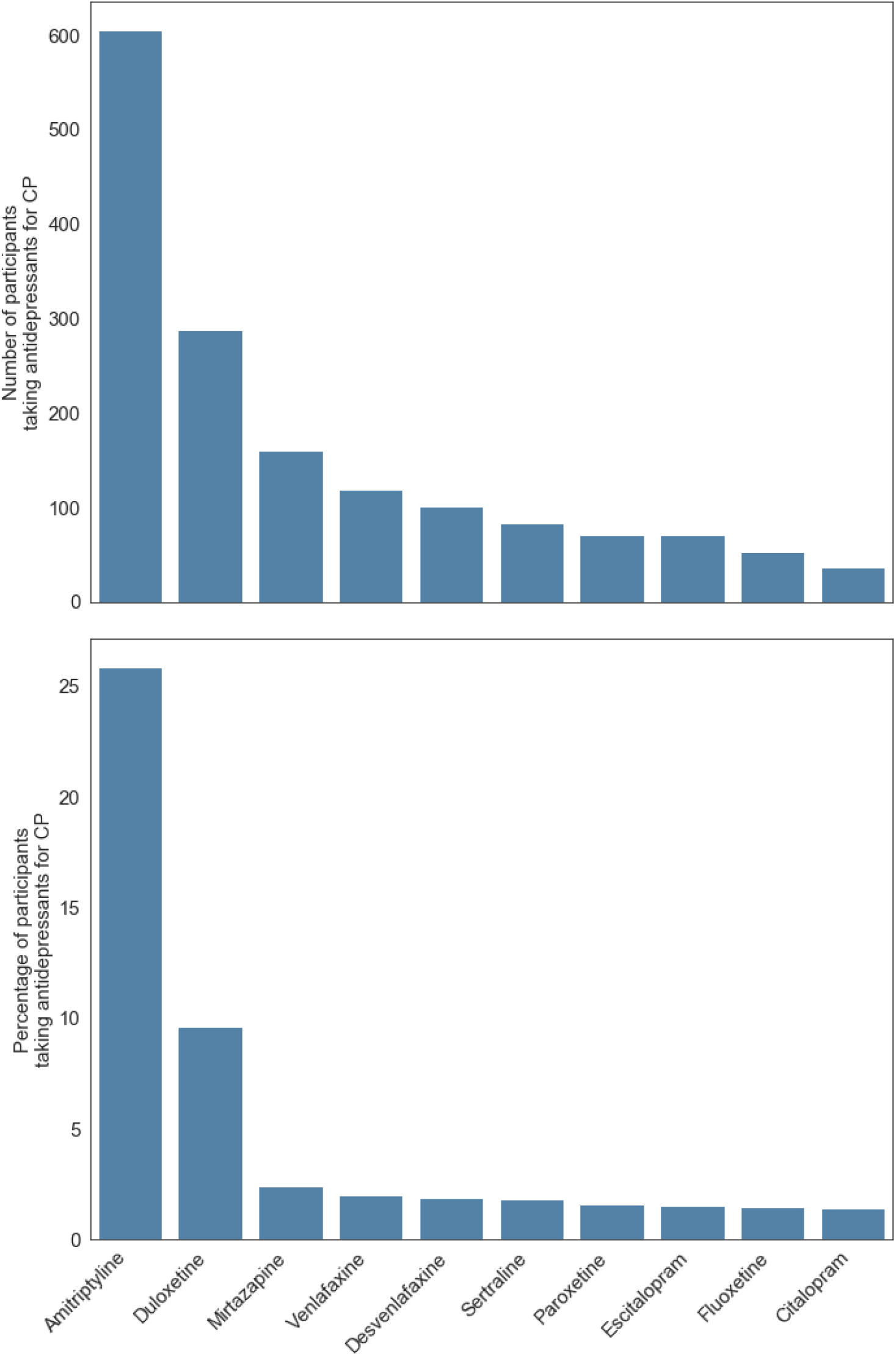
Antidepressant intake by chronic pain participants. Barplots showing the percentage (number) of participants with comorbid chronic pain (CP) and depression taking ten commonly used antidepressants in Australia (AGDS data only).

**Supplementary Figure 6.**
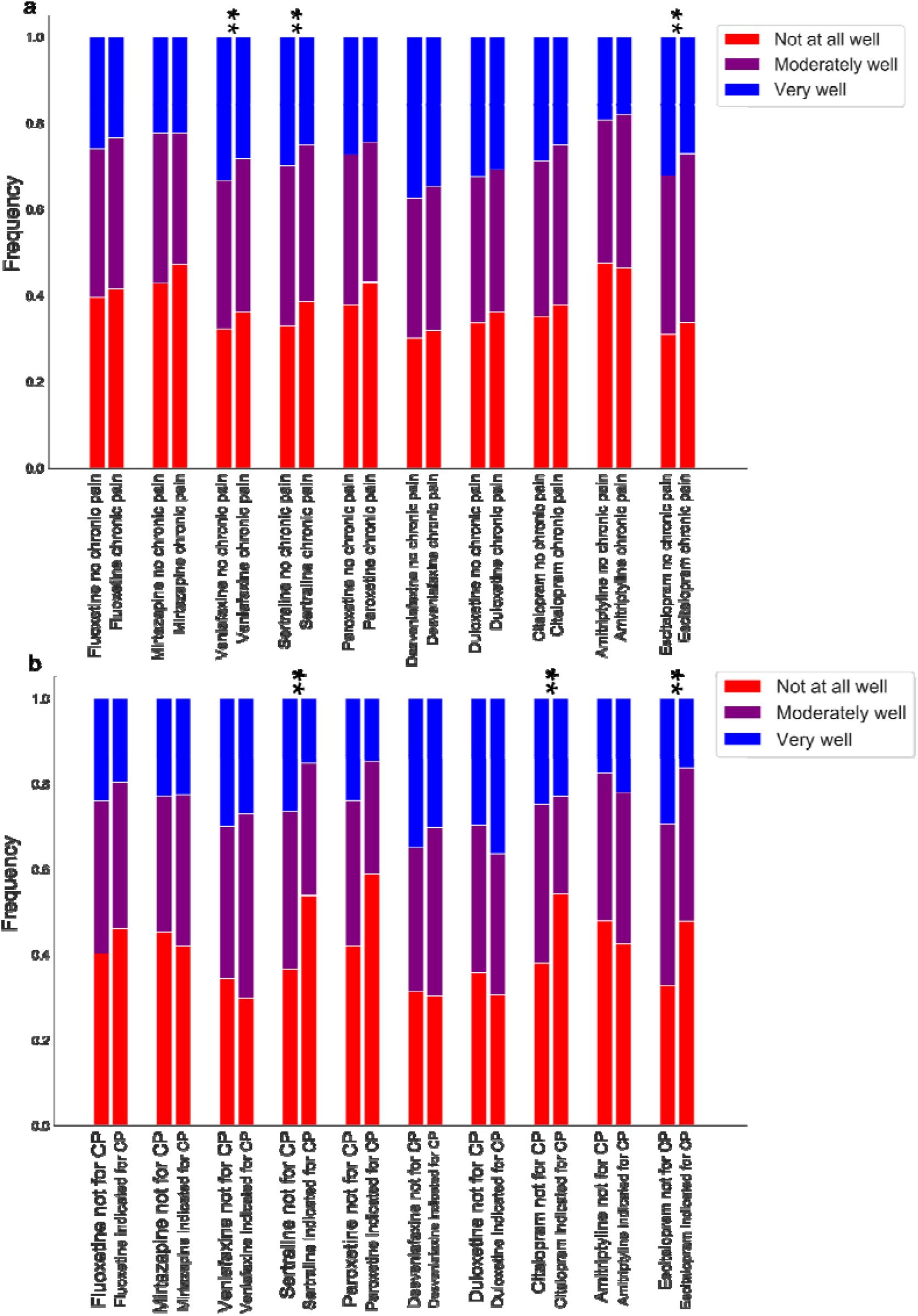
Association between antidepressant response, participants with chronic pain and antidepressant prescription for chronic pain. Bar plots depicting the average antidepressant response score across antidepressants stratified by participants reporting chronic pain (CP), regardless of: **(a)** the clinical indication for the antidepressant: or **(b)** whether the antidepressant was prescribed for chronic pain. Further details including p-values from a cumulative link logistic regression are in **Supplementary Tables 4 and 5** (AGDS data only).

## SUPPLEMENTARY TABLES

**Supplementary Table 1.** **Effect of demographic variables, depression and cohort on chronic pain**

|  |  |  |
| --- | --- | --- |
|  | OR (95%CI) | p-value |
| Cohort (AGDS) | 1.31 (0.96–1.77) | 0.086 |
| Sex (female) | 1.16 (1.07–1.26) | 0.0003 |
| Depression* | 1.95 (1.42–2.66) | 2.80E-05 |
| Education | 0.887 (0.86–0.91) | 1.00E-14 |
| Age | 1.02 (1.02–1.03) | 6.90E-74 |
| These results are from a multivariate logistic regression on chronic pain. Fully adjusted ORs are shown. Cohort represents a categorical variable of whether participants were from the AGDS cohort compared to the PISA cohort. *The effect of self-reported depression available on PISA and AGDS (all cases). Note the lack of association with cohort after adjustment for sex, age, education and depression. |  |  |

**Supplementary Table 2.** **Other psychiatric comorbidities with chronic**

| Diagnosis | OR (95% CI) | p-value |
| --- | --- | --- |
| Bipolar disorder | 0.947 (0.82–1.09) | 0.4570 |
| Premenstrual dysphoria | 1.19 (0.90–1.57) | 0.2355 |
| Schizophrenia | 1.58 (0.97–2.58) | 0.0675 |
| Anorexia nervosa | 0.788 (0.62–0.99) | 0.0443 |
| Bulimia | 1.17 (0.92–1.49) | 0.2003 |
| ADD/ADHD | 0.826 (0.68–1.01) | 0.0602 |
| Autism/Asperger | 1.14 (0.82–1.60) | 0.4294 |
| Tourette's | 1.12 (0.36–3.49) | 0.8502 |
| Generalized anxiety | 0.999 (0.92–1.09) | 0.9889 |
| Panic disorder | 1.02 (0.88–1.18) | 0.8050 |
| Obsessive-compulsive disorder | 0.945 (0.79–1.13) | 0.5344 |
| Hoarding disorder | 1.29 (0.72–2.32) | 0.3870 |
| PTSD | 0.987 (0.88–1.11) | 0.8335 |
| Phobia | 1.03 (0.82–1.29) | 0.7949 |
| Seasonal disorder | 0.885 (0.70–1.12) | 0.3179 |
| Social anxiety | 1.19 (1.04–1.36) | 0.0096 |
| Agoraphobia | 0.828 (0.63–1.09) | 0.1724 |
| Personality disorder | 1.11 (0.93–1.32) | 0.2440 |
| Substance use | 0.984 (0.79–1.22) | 0.8814 |
| Multivariate logistic regression assessing chronic pain comorbidities.<br>Multiple testing corrected significance threshold: $p < 0.0026$ | | |

**Supplementary Table 3.** **Correlations between recent substance use and chronic pain (CP)**

| Substance | CP OR (95%CI) | p-value |
| --- | --- | --- |
| GHB (gamma hydroxybutyrate) | 0.409 (0.14–1.21) | 0.105 |
| Inhalants | 0.723 (0.54–0.98) | 0.035 |
| Cocaine | 0.811 (0.65–1.01) | 0.060 |
| Ecstasy | 0.857 (0.69–1.06) | 0.161 |
| Other party drugs | 0.886 (0.54–1.46) | 0.632 |
| Stimulants | 0.986 (0.91–1.07) | 0.744 |
| Alcohol | 0.997 (0.99–1.00) | 3.17E-04 |
| Ketamine | 1.02 (0.71–1.45) | 0.933 |
| Hallucinogens | 1.03 (0.76–1.40) | 0.850 |
| E-cigarettes | 1.03 (0.95–1.12) | 0.421 |
| Amphetamines | 1.05 (0.86–1.28) | 0.640 |
| Sedatives or sleeping pills | 1.05 (0.99–1.11) | 0.080 |
| Tobacco | 1.05 (1.02–1.09) | 0.002 |
| Cannabis | 1.07 (1.01–1.13) | 0.016 |
| Painkillers or analgesics* | 1.29 (1.24–1.35) | 1.00E-29 |
| Opioids* | 1.31 (1.06–1.62) | 0.012 |

Alcohol use was measured as the number of days drinking three or more standard drinks in the past three months. All other variables were measured on a scale ranging from never to daily or almost daily for the past three months. \*Non-medical use of opioids & over-the-counter or other painkillers/analgesics

**Supplementary Table 4.** **Percentage of chronic pain participants with an antidepressant indication for chronic pain or for depression**

|  |  |  |
| --- | --- | --- |
| **Antidepressant** | **Chronic pain indication %** | **Depression indication %** |
| Sertraline | 2.83 | 97.29 |
| Escitalopram | 2.62 | 96.45 |
| Venlafaxine | 3.28 | 96.97 |
| Amitriptyline | 31.05 | 61.60 |
| Mirtazapine | 4.37 | 89.78 |
| Desvenlafaxine | 3.31 | 96.96 |
| Citalopram | 2.18 | 95.45 |
| Fluoxetine | 2.46 | 95.35 |
| Duloxetine | 15.32 | 94.95 |
| Paroxetine | 2.11 | 92.67 |

**Supplementary Table 5.** **Functional benefits associated with antidepressant use in participants with chronic pain**

|  |  |  |
| --- | --- | --- |
| **Self-reported benefit** | **OR (95% C.I.)** | **p-value** |
| Relief of depressive symptoms | 0.78 (0.71–0.87) | 1.5E-06 |
| Return of normal emotions | 0.80 (0.74–0.86) | 2.6E-08 |
| Getting back to normal daily activities | 0.82 (0.76–0.89) | 9.2E-07 |
| Improved relationships with those I am close to | 0.90 (0.83–0.97) | 0.01 |
| Restored control over my mood and actions | 0.90 (0.83–0.98) | 0.01 |
| Relief of other key symptoms | 0.94 (0.87–1.02) | 0.15 |
| Reduction in suicidal thinking or actions | 1.09 (1.01–1.18) | 0.03 |

**Supplementary Table 6.** **Effect of chronic pain on antidepressant response**

|  |  |  |  |  |
| --- | --- | --- | --- | --- |
| **Antidepressant** | **N** | **beta** | **S.E.** | **p-value** |
| Sertraline* | 5,189 | -0.29132 | 0.053 | 4.42E-08 |
| Escitalopram* | 3,877 | -0.28702 | 0.061 | 2.96E-06 |
| Venlafaxine* | 3,540 | -0.25168 | 0.065 | 1.13E-04 |
| Amitriptyline | 1,293 | -0.06086 | 0.134 | 0.650 |
| Mirtazapine | 1,718 | -0.16407 | 9.48E-02 | 0.083 |
| Desvenlafaxine | 2,262 | -0.14898 | 0.080 | 0.063 |
| Citalopram | 2,236 | -0.19203 | 0.082 | 0.019 |
| Fluoxetine | 3,178 | -0.1739 | 0.069 | 0.012 |
| Duloxetine | 1,776 | -0.18966 | 0.093 | 0.042 |
| Paroxetine | 1,361 | -0.30056 | 0.107 | 0.005 |
| *p<0.05 after Bonferroni multiple-testing correction for 10 variables/antidepressants |  |  |  |  |

**Supplementary Table 7.** **Effect of chronic pain indication on antidepressant treatment response**

|  |  |  |  |  |
| --- | --- | --- | --- | --- |
| **Antidepressant** | **N** | **beta** | **S.E.** | **p-value** |
| Sertraline* | 5,189 | -0.80599 | 0.207 | 1.00E-04 |
| Escitalopram* | 3,877 | -0.79848 | 0.253 | 0.002 |
| Venlafaxine | 3,540 | 0.143459 | 0.214 | 0.503 |
| Amitriptyline | 1,293 | 0.220429 | 0.119 | 0.064 |
| Mirtazapine | 1,718 | 0.003402 | 0.271 | 0.990 |
| Desvenlafaxine | 2,262 | -0.39922 | 0.270 | 0.140 |
| Citalopram* | 2,236 | -1.14947 | 0.379 | 0.002 |
| Fluoxetine | 3,178 | -0.40595 | 0.268 | 0.130 |
| Duloxetine | 1,776 | 0.2071 | 0.148 | 0.161 |
| Paroxetine | 1,361 | -0.50542 | 0.503 | 0.315 |
| *p<0.05 after Bonferroni multiple-testing correction for 10 variables/antidepressants |  |  |  |  |

